# Tract-based disconnection induced by WMH in patients with minor stroke is related to long-term post-stroke cognition

**DOI:** 10.1101/2023.08.04.23293684

**Authors:** Renaud Lopes, Grégory Kuchcinski, Thibaut Dondaine, Loïc Duron, Anne-Marie Mendyk, Hilde Hénon, Charlotte Cordonnier, Jean-Pierre Pruvo, Régis Bordet, Xavier Leclerc

## Abstract

**Background:** Over a third of minor stroke patients will suffer from post-stroke cognitive impairment (PSCI) but there are no validated tools to clearly identify at-risk patients in the early phase. We aimed to investigate the short and long-term cognitive decline using disconnection features from infarct and white matter hyperintensities (WMH) in first-ever minor ischemic stroke patients.

**Methods:** First-ever minor ischemic stroke patients (NIHSS≤7) were prospectively followed-up at 72-hour, 6- and 36-month post-stroke with cognitive tests and conventional brain MRI. Infarct and WMH volumes were semi-automatically evaluated on DWI and FLAIR sequences respectively. Bayesian models using tract-based structural disconnection were used to estimate the remote pathological effects of cerebral infarct and/or WMH. The disconnection approach was compared to features extracted from cerebral infarct and WMH volumes and locations. All lesion-based features were compared between patients with and without cognitive impairment at 6- and 36-month post-stroke. The potential association between the features and cognitive domains alterations was assessed by canonical correlation analyses. All statistical analyses were corrected for age, education and multiple comparisons.

**Results:** 105 patients (female, 31%) with a mean (± SD) age of 63 ± 12 years were enrolled. Infarct volume was 10.28 ± 17.10 cm3 and involved the middle cerebral artery territory in 83% of patients. The burden of WMH was higher within frontal periventricular white matter. Infarct-based features showed no significant relationship with 6 and 36-month PSCI. However, a WMH disconnection factor involving the commissural and frontal tracts was associated with 6- and 36-month PSCI, particularly in executive/attention, language and visuospatial domains. Memory domain alterations were associated with higher WMH burden in right temporal regions.

**Conclusions:** WMH-induced disconnectivity may predict short and long-term PSCI in minor ischemic stroke. These neuroimaging features extracted from routine MR sequences could help identifying at-risk patients to test future rehabilitation interventions.

## Introduction

Over a third of stroke patients will suffer from post-stroke cognitive impairment (PSCI) but there are no validated tools to clearly identify at-risk patients in the early phase. PSCI are influenced by multiple factors including age, stroke recurrence and preexisting brain injury ^1^. Infarct location and volume also play a role in PSCI, considering strokes of various severities^2^. Moreover, secondary alterations depending on the initial infarct location are related to long-term cognitive and functional outcomes supporting the hypothesis neurodegeneration due to infarct-induced disconnection ^3^.White matter hyperintensities (WMH) are frequently observed on brain MRI in stroke patients and constitute an independent risk factor in the development of cognitive disorders ^4^. Neuropathology studies showed that WMH visible on MRI reflect neurodegeneration and axonal loss ^5, 6^. These alterations of white matter tracts are likely to impact connected structures through anterograde and retrograde degeneration phenomena ^7^. However, the impact of infarct and WMH on the long-term cognitive outcome is much less established in minor stroke, despite a prevalence of PSCI ranging from 30 to 55% in this specific context ^8, 9^.

Generally, secondary alterations induced by infarct and WMH are measured using MRI sequences not acquired in clinical practice, such as brain atrophy on 3D T1-weighted images ^7^, iron load on quantitative susceptibility images ^7^, functional connectivity on resting-state fMRI ^10^ and structural connectivity on diffusion tensor imaging ^11^. In this study, we are interested in MRI features extracted from clinically accessible MRI sequences. Presumed secondary alterations were quantified by analyzing white matter fiber bundles directly involved by infarct or WMH through the concept of structural connectivity ^12^. The development of large databases of diffusion MRI data and the analysis of structural connections among brain regions that are common in a large group of healthy subjects have allowed the construction of so-called connectome and tractographic atlases ^13^. Disconnectome approach estimates structural disconnection from lesion mask and tractographic atlas, by measuring the probability of normal white matter tracts passing through the lesion ^14^. Approaches using white matter tract disconnection from infarct lesion have been used to investigate brain network dysfunction, behavioral and cognitive deficits after stroke ^15, 16^. No study has investigated the relationships between PSCI and white matter tract disconnection induced by WMH.

Infarct and WMH can alter multiple white matter tracts and are associated with cognitive impairment in different and/or multiple domains. Thus, the interaction between WMH and acute ischemic event can trigger a series of pathological events that lead to heterogenous trajectories of cognitive decline ^17^. To address this heterogeneity, dimensional methods such as factor analysis^18^ can be employed to identify patterns of observable and latent anomalies at the patient level. This approach enables the possibility that multiple latent factors of white matter tracts disconnection can be expressed to varying degrees within a patient.

The objective of this study was to investigate the associated factors with the short- and long-term cognitive decline using Latent Dirichlet Allocation (LDA) to identify multiple latent disconnection factor from infarct and WMH lesions in first-ever minor ischemic stroke patients. We compared the approach with global and regional analyses of the lesions volume and location. We hypothesized that PSCI could be explained by a combination of white matter disconnected tracts by infarct and/or WMH, which could account for the variations in multidomain cognitive decline among stroke patients.

## Methods

### Study population

Between 2010 and 2020, a total of 202 patients with first-ever ischemic stroke were enrolled in the Study of Factors Influencing Poststroke Dementia cohort (STROKDEM; NCT01330160). Participants aged 18 years and older were included, while patients with the following criteria were excluded: (1) prestroke dementia (defined as a short form Informant Questionnaire of Cognitive Decline in the Elderly [IQCODE] score of 64 or more) ^19^, (2) moderate or severe stroke (NIH Stroke Scale [NIHSS] score > 7), (3) no MRI-visible lesion, (4) secondary hemorrhage, with type 2 parenchymal hematoma ^20^, (5) MRI contraindications, (6) inability to speak, and (7) a poor understanding of the French language. The STROKDEM study conducted follow-up assessments of the participants’ clinical, neuropsychological, and imaging from 72 hours to 36 months post-stroke.

### Neuropsychological Assessment

The participants’ cognitive functions, including executive functions/attention, memory, language, and visuospatial abilities, were assessed 6- and 36-month post-stroke using a battery of neuropsychological tests (T.D.) (see Supplemental file). For each participant with complete data, test-specific z scores based on available norms adjusted for age, sex, and education were calculated and averaged by domain to obtain summary domain-specific z scores. Each participant’s cognitive status was reviewed in a multidisciplinary staff meeting at 6- and 36-month post-stroke, based on any complaints and symptoms during clinical examination, and the neuropsychological test results.

### MRI acquisitions

All participants underwent structural brain MRI within 72 hours of admission to the hospital using one 3T MRI scanner (Achieva, Philips, Best, the Netherlands). The imaging protocol notably included a 3D T1-weighted (T1W) gradient echo sequence, a diffusion-weighted imaging (DWI) sequence, and a fluid-attenuated inversion recovery (FLAIR) sequence (see Supplemental Material for sequence parameters).

### Infarct- and WMH-based features

Normalized volumes of infarct and WMH were calculated. A vascular territory template in MNI space was used to create an incidence map of infarct location using 6 supratentorial and 4 infratentorial regions^21^. Normalized WMH volumes were calculated in 36 distinct parcels^22^ (see Supplemental Material for more details).

### White matter tract-based disconnection

To estimate how focal lesions may lead to pathological effects distally from the primary site of lesions, we used a tract-based structural disconnection approach ^14^. A publicly available diffusion MRI streamline tractography atlas, constructed using data from 842 Human Connectome project participants, was used ^13^. The corpus callosum tract was subsequently divided into five segments to enhance the interpretability of callosal disconnection, resulting in a total of 70 tracts ^15^. For each tract, the number of streamlines passing through the infarct or WMH mask (« disconnected streamlines ») was converted to a percentage of the total number of streamlines assigned to that tract in the tractography atlas. The approach was independently repeated for infarct and WMH masks, resulting in an estimate of percent disconnection severity for each tract and both masks.

### Modelling latent disconnection factors

Since we hypothesized that cognitive impairment in several domains could be linked to a combination of « disconnected » tracts, we used a hierarchical Bayesian model, called Latent Dirichlet Allocation (LDA), that allows each patient to express one or more latent disconnection factors, each of which is associated with distinct but possibly overlapping tract-based disconnection patterns ^23^. The percentage of disconnected streamlines for each tract was considered as input of the LDA model. Given a user-defined number of factors K, a variational expectation-maximization algorithm was applied to estimate the probability of an individual expressing a latent factor or factor loading [Pr(Factor │ Participant)] and the probability that a factor was associated with disconnection at a tract [Pr(Tract │ Factor)]. The final K for subsequent analyses was determined by choosing the one that offered the highest stability across runs ^23^ (see Supplemental file).

### Comparing cognitive status across latent factors

Quade’s nonparametric analyze of covariance and pairwise comparisons among groups were applied to explore how the local and regional volumes and the factor loadings, i.e., Pr(Factor │ Participant), varied across 6 and 36 months post-stroke cognitive status. Two analyses were performed. First, participants were divided into two groups based on the presence or not of 6-month PSCI established by the multidisciplinary staff. Second, four groups were defined based on the trajectory of their cognitive status between 6- and 36-month post-stroke: converters were Conv-Cog+ (new-onset cognitive impairment at 36 months) and Conv-Cog-(cognitive impairment at 6 months but not at 36 months), whereas non-converters were Noconv-Cog-(no cognitive impairment at both time points), and Noconv-Cog+ (cognitive impairment at both time points). As age and educational level differed across groups, they were used as covariates in all statistical analyses and false discovery rate (FDR) was used to correct for multiple comparisons with a statistical significance level set to p < 0.05.

### Correlations with cognitive performance

To find an optimal linear combination of the four cognitive domains z scores at 6- and 36-month post-stroke that maximally correlated with the lesion-based features, Canonical Correlation Analysis (CCA) was applied between the cognitive z scores and each feature, and repeated at 6- and 36-month post-stroke. Age and educational level were used as covariates. Permutation test and multiple comparisons corrected using FDR were used to assess the statistical significance of CCA (p<0.05).

### Standard Protocol Approvals, Registrations, and Patient Consents

STROKDEM study was conducted in accordance with the tenets of the Declaration of Helsinki and was approved by local institutional review boards. Prior written informed consent was obtained by all patients or their legal representatives (Comité de Protection des Personnes Nord Ouest IV, Lille, France; reference: 2009-A00141-56, March 17, 2009).

### Data availability

The results can be made available on reasonable request and the data have been shared with the STROKOG and METACOHORT consortia ^24, 25^.

## Results

### Demographic, clinical, and imaging characteristics at baseline

Out of the 202 eligible patients, 178 met inclusion criteria, and 105 underwent evaluations at both 6- and 36-month post-stroke and were included in the current analysis (mean age 63 ± 12; mean educational level 12 years; 31% female) (Figure S1). There were no significant differences between those who did and did not complete the follow-up evaluations (Table S1). On admission, the study participants had a median NIH Stroke Scale (NIHSS) score of 1 and a mean infarct volume of 10.28 ± 17.10 cm^3^. None of the participants experienced a second stroke during the study.

### Changes in cognitive status between 6- and 36-month post-stroke

Six months after stroke, 55 participants (52%) had PSCI. They were older (p = 0.038) and had a lower educational level (p=0.036) than participants without PSCI (Table S2).

Thirty-six months after stroke, the Conv-Cog+, Noconv-Cog−, Noconv-Cog+, and Conv-Cog− contained 10, 40, 30, and 25 participants, respectively. Hence, there were 35 converters (33%). The 4 groups differed according to the educational level, with a lower educational level for the Noconv-Cog+ group vs the Noconv-Cog− group (Table S2).

In the following results, age and educational level were used as covariates in statistical analyses.

### Associations between infarct- or WMH-based features and cognitive impairment at 6- and 36-month post-stroke

Infarct volume and territory, and WMH volumes were not associated with 6- and 36-month PSCI (Table S1). While WMH burden tended to have the same distribution in participants with and without 6-month PSCI, participants in Conv-Cog+ group at 36 months showed higher WMH burden in the 3 layers closest to the ventricle of the right temporal lobe compared to the other groups (Figure 1-C).

**Figure 1:**
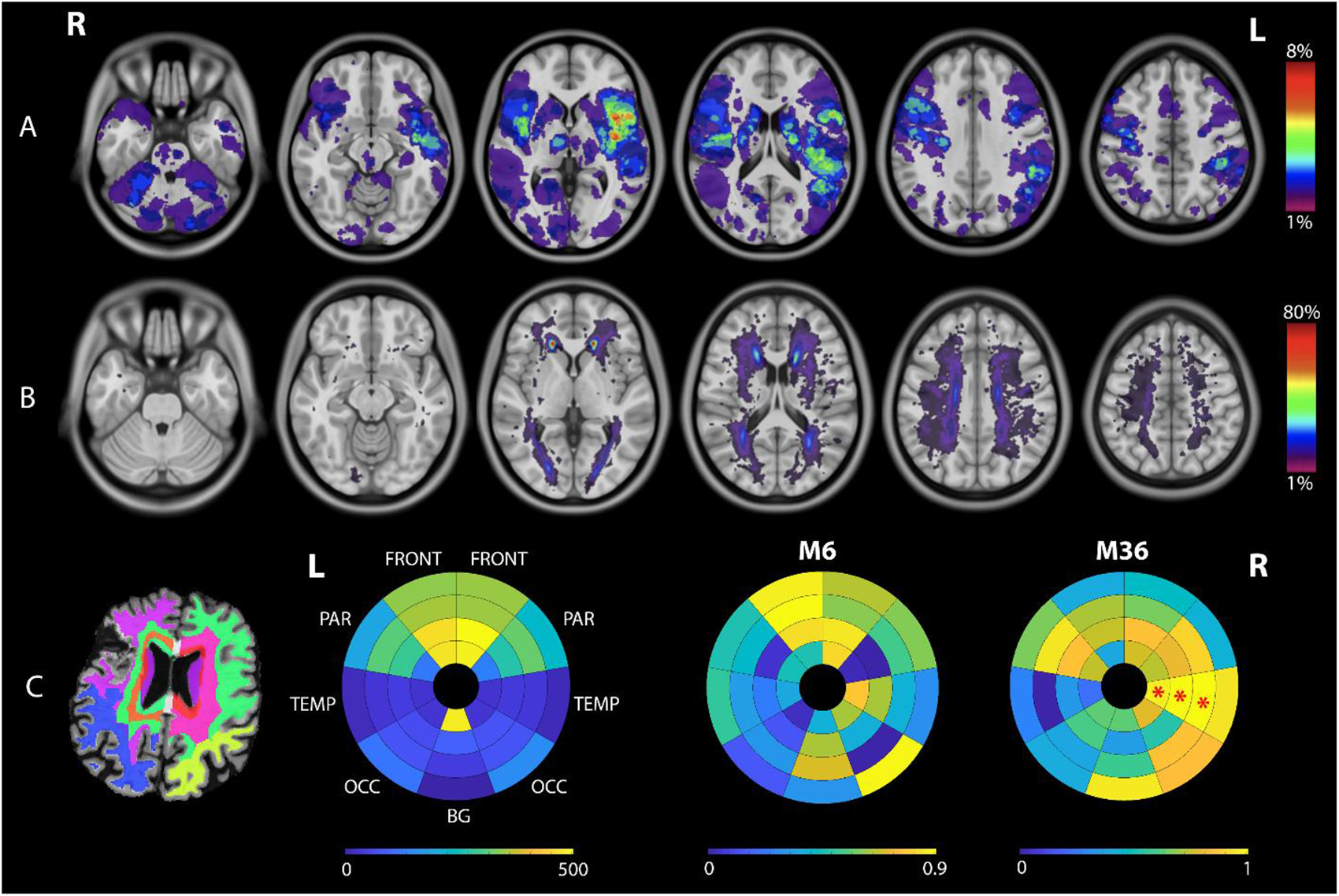
Topography of the infarcts (A) and WMH (B), with a lesion overlay map atlas. (C) Regional patterns of WMH. From left to right: an example of lobar segmentations, bullseye representation of WMH load for all participants, bullseye representation of the statistical difference between participants with and without 6 months PSCI (color scale: 1 – p-value), bullseye representation of the statistical difference between the 4 groups at 36 months PSCI (color scale: 1 – p-value). Color scale for (A) and (B): number of participants with lesions at each voxel. * p < 0.05 FDR corrected BG basal ganglia, FRONT frontal lobe, PAR parietal lobe, OCC occipital lobe, TEMP temporal lobe

The Bayesian model was applied to infarct and WMH disconnected tracts with a number of latent factors, K = 2 to 10. For both Bayesian models, the 3-factor model was the most stable.

The Bayesian model identified three latent factors using disconnected tracts by infarct that differed with respect to hemisphere and infra-tentorial signatures (Figure 2-A). Factor 1 was defined almost exclusively by disconnections of right hemisphere tracts. The ten most severe disconnected tracts accounted for 66% of the disconnection load (i.e. the summation of posterior probability across tracts), defined by 4 projection, 4 association and 2 brainstem tracts. Factor 2 was mostly the symmetric of factor 1 for the left hemisphere. The ten most severely disconnected tracts accounted for 65% of the disconnection load, defined by 4 projection and 6 association tracts. Factor 3 was mainly defined by disconnections of infra-tentorial tracts. The ten most severe disconnected tracts accounted for 75% of the disconnection load, defined by 3 association, 3 brainstem, 3 cerebellum and 1 commissural tracts. Factor loadings were similar between cognitive status groups at 6- and 36-month post-stroke (Table 1).

**Figure 2:**
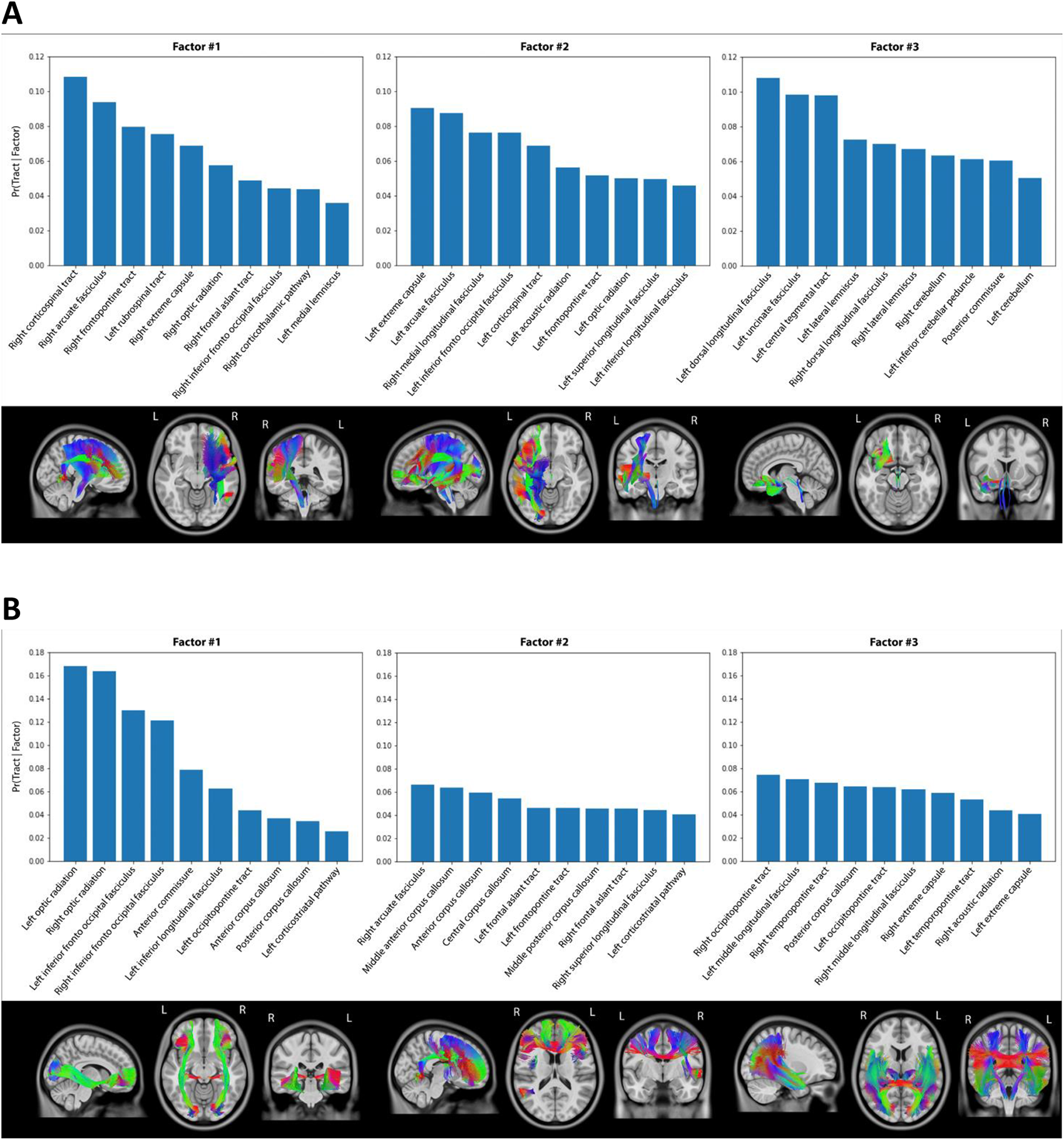
The 3 latent tract-based disconnection factor estimated from (A) infarcts and (B) WMH of participants. The first line displays the probability of disconnection at that tract for a particular factor (i.e., Pr(Tract | Factor). The second line displays the 5 tracts with the highest probability of disconnection.

**Table 1.**
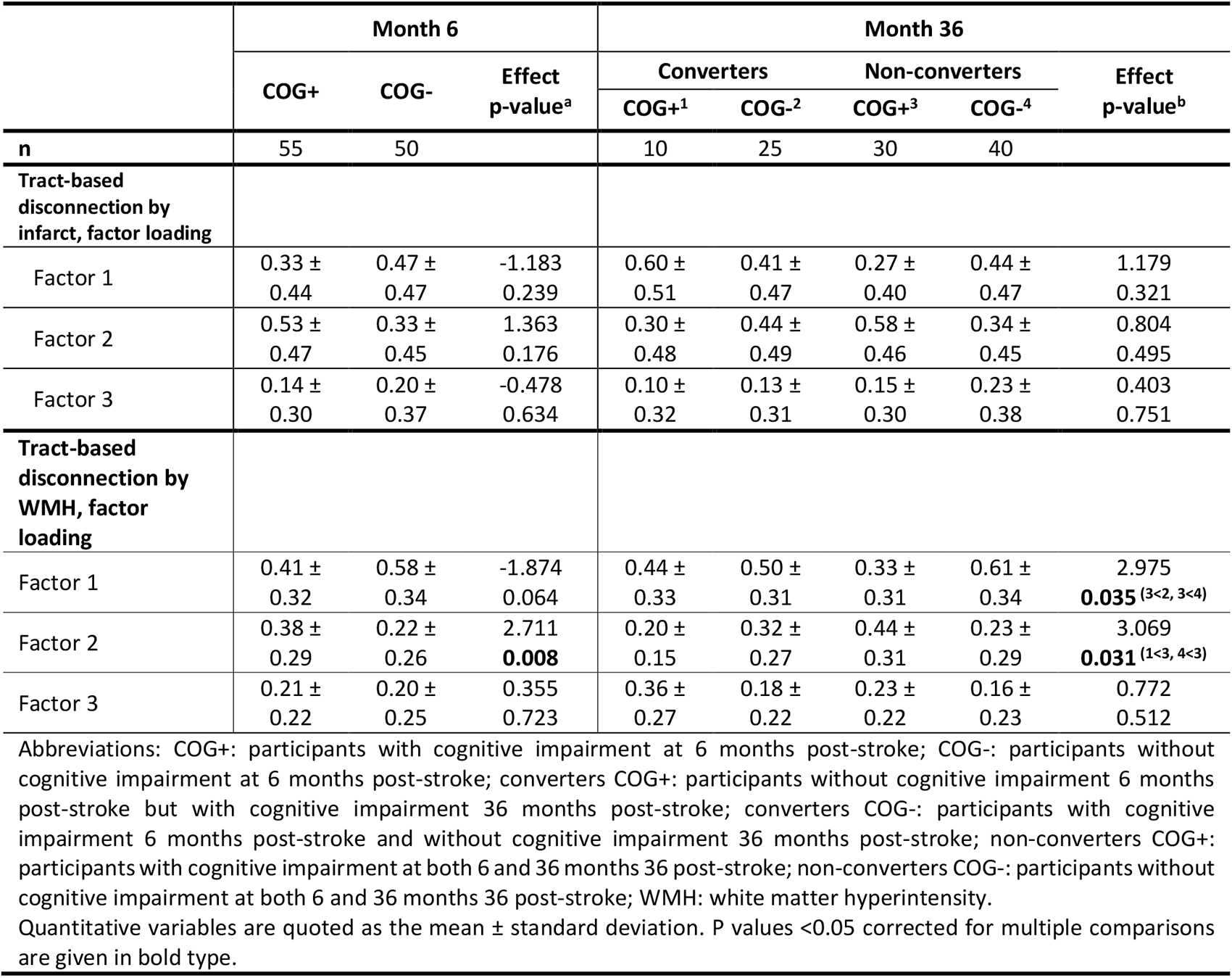
Associations between tract-based disconnection factors and cognitive status at 6 and 36 months poststroke.

By using disconnected tracts by WMH, the Bayesian model identified three latent factors that differed with respect to commissural and disconnection load (Figure 2-B). For factor 1, more than half of the disconnection load was defined by two tracts (optic radiation and inferior fronto-occipital fasciculus). Factor 2 was mainly dominated by disconnections of commissural and frontal tracts. The ten most severe disconnected tracts accounted for 51% of the disconnection load, defined by 4 commissural, 4 association and 2 projection tracts. Factor 3 was mainly characterized by disconnections of temporo-occipital tracts. The ten most severe disconnected tracts accounted for 60% of the disconnection load, defined by 5 projection, 4 association and 1 commissural tracts. Factor 2 loading was higher for participants with 6 months PSCI (Table 1). At 36-month post-stroke, participants in Noconv-Cog+ group had higher factor 2 loading than participants in Conv-Cog+ and Noconv-Cog-groups.

### Associations between significant markers of PSCI and cognitive domains at 6- and 36-month post-stroke

To examine associations between previous significant markers and cognitive domains, CCA was performed between each marker and 4 cognitive domain-specific z scores. Factor 2 from WMH-induced disconnection was associated with worse impairment in language and attention domains at 6-month post-stroke (Figure 3-A) and with worse impairment in attention, language and visuospatial domains at 36-month post-stroke (Figure 3-B). Higher WMH burden in the second layer closest to the ventricle of the right temporal lobe was associated with worse impairment in memory domain (Figure 3-E).

**Figure 3:**
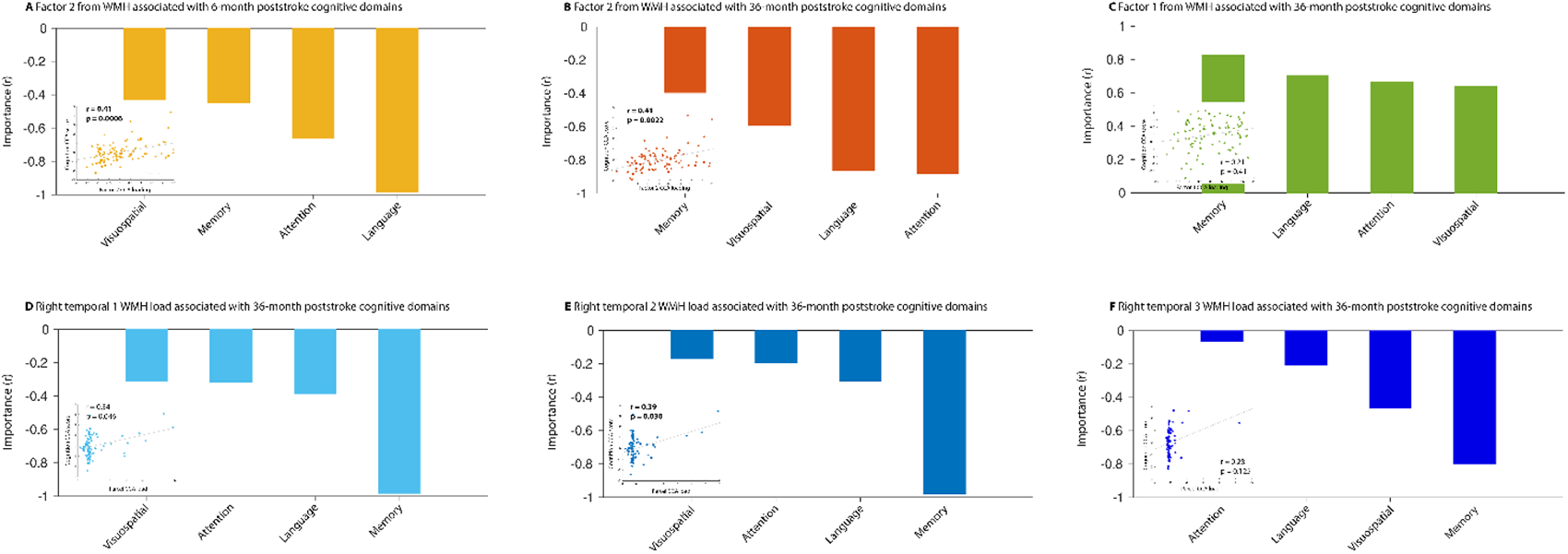
Canonical correlation analyses between the 4 cognitive domain z scores and the significant lesion-based features from the comparisons of 6 and 36 months PSCI. The first line displays the results between the loadings of factors identified from disconnection by WMH and 4 groups of cognitive z score. The second line displays the results between the normalized WMH load in parcels and 4 groups of cognitive z score. Positive correlation suggests that a higher loading was associated with greater impairment. The scatter plots show the relationships between CCA cognitive score and CCA loading, where each dot represents a participant. For example, Factor 2 was associated with worse visuospatial, memory, attention and language functions 6 months poststroke.

## Discussion

In this study, we found no relationship between infarct-based features and 6 and 36-month PSCI whereas a disconnection factor induced by WMH was found in the commissural and frontal tracts and was associated with 6- and 36-month PSCI, especially in executive/attention, language and visuospatial domains. These findings suggest that in minor strokes, ongoing underlying chronic cerebrovascular lesions already present at stroke onset are related to long-term PSCI.

Patients with minor stroke have higher risk of cognitive impairment and even dementia than the general population ^26, 27^. Understanding the underlying mechanisms of cognitive impairments following a minor stroke is challenging due to the size of the stroke. Indeed, infarct volume and location are common markers of PSCI ^2^. However, the effects are more pronounced for severe strokes than for minor strokes ^26^. Additionally, the approach used, which relies on voxel-based lesion-symptom mapping, requires a sufficient brain overlap of infarcts to perform statistical comparisons. Given the small sizes of infarcts in our population and the limited number of subjects, there was a low overlap between lesions. Therefore, we resorted to alternative approaches.

In this study, we developed a disconnection-symptom mapping technique to address the relatively small sample size and to generate predictions regarding the remote white matter bundles where degeneration is likely to occur. One strength of this approach is the requirement of only conventional MR sequences. Given that the heterogeneity in the rate of progression of PSCI and the involvement of multiple cognitive domains ^17, 36^, we used a data-driven Bayesian framework, called latent Dirichlet allocation (LDA), to model the possibility that multiple latent factors are expressed to varying degrees within a subject.

As expected, the infarct volume and location were not associated with PSCI in our population. Although the disconnection approach allowed us to statistically assess the infarct’s impact on PSCI, the lack of association prevented us from drawing definitive conclusions regarding the infarct’s effects on patients with minor strokes. The low overlap may also explain the limited interest of the Bayesian model based on the disconnection of white matter tracts by the infarct. The three-factor model identified three different disconnection topographies related to the damaged vascular territory. Indeed two factors were symmetric and represented patients with stroke in left or right supra-tentorial stroke, and the last factor represented patients with infra-tentorial stroke. As a result, the Bayesian model’s assumption that patients could express several multiple disconnected tract factors was limited.

In our study, PCSI was associated with WMH-based features. WMH load is considered as a correlate of long-term cognitive impairment ^30, 31^. However, we did not find any association between total WMH volume and 6- or 36-month PSCI when adjusted for age and education level. A previous study on participants with minor stroke showed a relationship between the total WMH load and persistent cognitive impairment (>30 days), but WMH volume no longer predicted PSCI when adjusted for age ^4^. Older age and lower levels of education are consistent risk factors for PSCI ^32^. Our results were consistent with this statement as participants with 6-month PSCI had higher WMH volume than those without 6-month PSCI when adjusted only for education level (p=0.035).

Tract-based disconnection induced by WMH was the most predictive marker of long-term PSCI, supporting the assumption that WMH affect cortical connectivity by diminishing efficiency of neural transmission, resulting in cognitive impairment ^33^. One latent factor (Factor 2) composed of commissural and frontal tracts was associated with cognitive domains at 6 and 36 months after stroke, and more specifically with attention and language domains 6-month post-stroke and with visuospatial, language and attention 36-month post-stroke. Our results are consistent with a recent study using the same dataset showing a relationship between functional connectivity from frontal regions at 6 months post-stroke and cognitive z scores of attention and visuospatial domains at 36 months post-stroke ^9^. Vascular lesions within corpus callosum have been associated with worse performance on cognition and reduced microstructural white-matter integrity within both the corpus callosum and the whole-brain white matter ^34^. Moreover, Zhao et al. showed that WMH location for long-term PSCI was mainly in the corpus callosum ^35^. The integrity of commissural pathways are essential for selective attention as well as processing and integration of visuospatial information ^36^. Language impairments at 6- and 36-month post-stroke were also highly associated with the Factor 2 loading. The arcuate fasciculus and the frontal aslant tract were among the ten most severe disconnected tracts represented by Factor 2. The arcuate fasciculus has a major implication in language processing ^37^ by connecting the key speech production region in the frontal lobe with the speech comprehension region in the posterior temporal lobe ^38^. The frontal aslant tract connects the inferior frontal gyrus with the superior frontal gyrus and cingulate gyrus and sulcus. This tract has been found to be highly associated with speech initiation, verbal fluency, stuttering and executive functions ^39^. Lastly, our regional analysis of WMH load revealed that memory function impairments were mostly associated with WMH load in right temporal regions. However, further investigations are needed because the statistical significance was driven by a small proportion of patients with WMH load in temporal regions, all of whom had memory function impairments. Nonetheless, temporal regions are essential for memory encoding and retrieval, and higher WMH load contributes to verbal memory deficits ^40^.

Our study includes limitations. Although our tract-based structural disconnection approach is popular and validated, it is an indirect method to assess the degree of disconnection of each tract by assuming that all brains have the same white matter anatomy. Atlas-based approaches are not sensitive against potential biases arising from interindividual differences. Indeed, it is important to highlight that the white matter tract atlas used in our study was based on very high-quality data with 90 direction high-angular resolution diffusion imaging, and it was derived from a large sample of participants (N = 842 participants). While the atlas provides valuable information for our current analysis, a further study could explore the potential benefits of creating a white matter tract atlas specifically tailored to the demographics of the patient population under investigation. Secondly, the accuracy of tract anatomy may be affected by the relatively large voxel size of the DWI sequence (14.4 mm^3^) compared to the FLAIR sequence (1.6 mm^3^). Finally, as tract-based disconnection measures are implicitly related to the lesion, the utility of these approaches still needs to be demonstrated in very large cohorts where the information between voxel-based lesion and tract-based disconnection could be similar. However, even if the information overlaps, tract-based disconnection measures may be useful from a neuroscience perspective, notably in the investigation of brain network alterations.

## Conclusion

The present study identified a marker of tract-based disconnection information induced by WMH and associated with long-term PSCI in patients with minor strokes. Further studies on larger cohorts of patients are needed to confirm these findings. This marker can be easily implemented in clinical practice as it solely relies on conventional MR sequences, making it useful for selecting patients at high risk of post-stroke dementia. Such patients may benefit from early cognitive rehabilitation and personalized therapy.

## Data Availability

The results can be made available on reasonable request and the data have been shared with the STROKOG and METACOHORT consortia.

## Non-standard Abbreviations and Acronyms

PSCI: Post-stroke cognitive impairment WMH White matter hyperintensities
NIHSS: National Institutes of Health Stroke Scale MNI Montreal Neurological Institute
LDA: Latent Dirichlet Allocation
CCA: Canonical Correlation Analysis
Conv-Cog+: New-onset cognitive impairment at 36 months
Conv-Cog-: Cognitive impairment at 6 months but not at 36 months
Noconv-Cog-: No cognitive impairment at both time points
Noconv-Cog+: Cognitive impairment at both time points

## Supplemental Material

### Supplemental methods

Neuropsychological Assessment

MRI acquisitions

Infarct- and WMH-based features

Modelling latent disconnection factors

Selection of the number of latent factors

### Supplemental results

Figure S1

Table S1

Table S2

References18,21–23,41–47

